# Comparison of binary classifiers in forensic dentistry for sex determination

**DOI:** 10.1101/2025.05.07.25326932

**Authors:** Álvarez-Vaz Ramón, Sassi Carlos, Verónica Gargano, Picapedra Alicia

## Abstract

In forensic odontology, teeth are essential components of the stomatognathic system, since they constitute excellent material for research, thanks to their almost indestructible structure and therefore the information linked to their size and characteristics, is extremely useful in determining sex and age. In this study, which corresponds to a sample of 524 lower plaster models (286 male subjects and 238 females) of patients assisted in an orthodontic clinic in Montevideo, Uruguay, the mesiodistal diameter (width), and gingivo-incisal height of the 2 canines and the intercanine distance are measured for sex identification based on odontometric measurements and their ratios. Different binary classifiers from what is known as supervised classification methods are applied in the statistical learning paradigm. Several methods are proposed and their performance is evaluated by working with test and learning samples and evaluating different metrics to measure the accuracy of the models, showing a performance greater than 65%. The details of this project are on the OSF platform in the project.*Relaciones Odontométricas en Odontología Forense (Odontometric Relationships in Forensic Dentestry)* en https://osf.io/javru/

## Introduction

There has long been a need and desire for human identification, given their sociopolitical nature and ambition to differentiate themselves from others. Indeed, the dynamics and changing collective organizations are characterized by the desire to individualize and hold their members civilly, administratively, commercially, and criminally liable. [1], [2], [3]. In the forensic field, a positive identification must be based on the intervention of a multidisciplinary team [2, 4, 5] capable of performing an appropriate reconstruction of the individual’s biological profile, using four essential components: *age, sex, height, and ancestry*. [6], [7]. Dental organs exhibit taphonomic resistance, being composed of almost indestructible structures, thus enabling post-mortem odontometric, comparative, and reconstructive analyses. [8], [9]. The most commonly used measurements, obtained from plaster models, are the mesiodistal, vestibulopalatine, mesiovestibular-distopalatine, and distovestibular-mesiopalatine diameters, the gingivoincisal height, and the intercanine distance, which constitute several indices. [3, 6, 7, 9–13]. Teeth, in general, and permanent canines in particular, are indicated to demonstrate sexual dimorphism by several authors in different populations around the world, [14–16]. In these studies, taking size into account has guided the process of sex determination, taking into account the fact that most teeth develop before skeletal maturation, making these measurements a valuable indicator of sex, mainly in subadult individuals, in the absence or emergence of secondary sexual characteristics, [17]. Legal and/or forensic dentistry has benefited from the invaluable contribution of statistics, developing predictive models tailored to the conditions of each case, [7]. In view of the above, this work seeks to find methods that allow identifying the sex with the highest degree of accuracy, using statistical models, as parsimonious as possible, based on the measurements of lower canines, and the relationships between these, for a sample of plaster models of patients assisted in an orthodontic clinic in the city of Montevideo, Uruguay.

For this purpose, the work is divided into 3 parts, the first one presents some binary classifiers in the *Methodology* section that can be used for the previously proposed objective, then we see how they work empirically as shown in the *Materials* section and then in the *Conclusion* section we propose some alternatives to always improve the models for sex identification based on odontometric measurements.

## Methodology

There are several binary classifiers that can be used in the field of statistical learning, more specifically what is known as supervised classification, where there is an input data matrix *X* and one or more variables *Y* with labels. From *X*, each observation *x*_*i*_ is classified, assigning it a label. For this purpose, some binary classifiers are presented, which are based on different types of input variables.

### Discriminant Analysis Method

Discriminant Analysis (DA) is a multivariate statistical technique for supervised classification but has two purposes, the description, where it is of interest to analyze whether there are differences between a series of groups into which a population is divided with respect to a set of variables and, if so, to find out why. On the other hand, DA seeks to make predictions through a systematic procedure for classifying new observations of unknown origin into some of the groups considered, [18], [19], [20]. Although this work focuses on binary classifiers (i.e., there are two groups), there are *k* samples of size *n*_*g*_ (*g* = 1, 2, … *k*) from *k* populations from which *p* quantitative characteristics are measured. Using this information, we want to determine which of those *k* populations a new observation is most likely to have been randomly selected from. Each element of the population is assigned to one of the groups according to a specific decision rule, trying to make the smallest possible errors. The analysis has a certain ”predictive” power because, in a way, the criteria used to classify a current population can be used for new elements that are incorporated into it. The disadvantage of this method is that it is limited to quantitative explanatory variables that also follow a varied Normal distribution. Functions are sought that best discriminate between groups, which is why it is necessary to define decision rules, ensuring the fewest errors possible.

When classifying, three types of distance are of interest:

- Distance between units.
- Distance between populations.
- Distance between unit and population.

### Distance between individuals

The Mahalanobis distance between individuals *i* and *l* is:

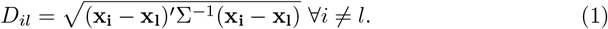

where 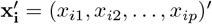 observed vector of individual *i*, Σ variance and covariance matrix.

### Distance between populations or groups

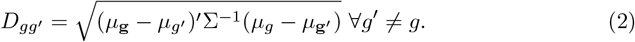

where 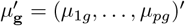 centroid, vector of means of the *p* variables. Σ matrix of variances and covariances.

### Distance between individual *i* and centroid of each group

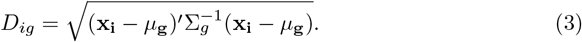

where 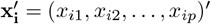 observed vector of individual *i*. Σ_*g*_ variance-covariance matrix of group *g*

Unit *i* is classified into group *g* if 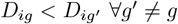.

In generic form, *P* (*g*^*′*^| *g*) is the error of classifying an observation from group *g* into group *g*^*′*^, where the probability of error of classifying an observation from group *g*^*′*^ into group *g* is:

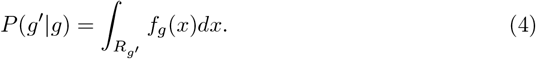

The probability of misclassifying all observations belonging to group *g*:

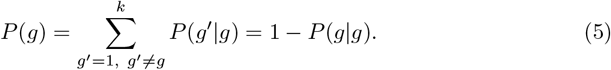

The total probability of misclassification is:

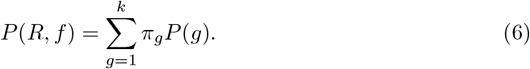

The decision rule is the one that minimizes the total probability of error.

### Principle of maximum likelihood

The principle consists in assigning observation *i* to the population where the observed vector 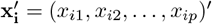 has the greatest likelihood of occurring, that is, *i* is assigned to group *g* if:

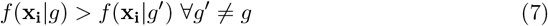

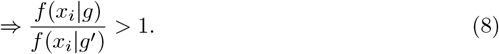

### Principle of a posteriori probability

The principle is to assign observation *i* to the population with the highest posterior probability (the probability that *i* belongs to *g* conditioned on the observed vector *x*_*i*_). The posterior probability, according to Bayes’ Theorem, is stated as follows:

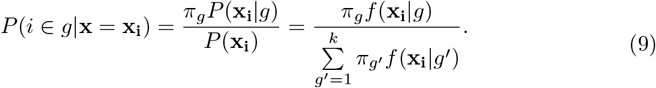

where *π*_*g*_ = *P* (*i* ∈ *g*) is the prior probability of belonging to group *g*. Unit *i* is classified into group *g* if:

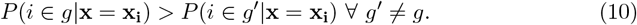

The proposed rules are optimal if the joint distribution is known, with all parameters known.

### Classification Rules applied to 2 groups

Given **x**_**i**_ ∼ **N**_*p*_(*µ*, Σ) the density function is:

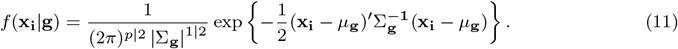

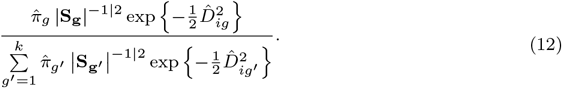

Two cases are distinguished:

1. In the case of equality of variance matrices, observation *i* is assigned to group *g* if:

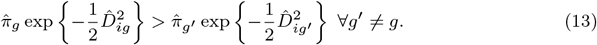
2. In the case of matrices of different variances, observation *i* is assigned to group *g* if:

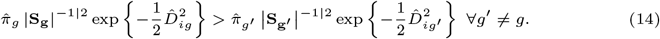 When the cost of error is not considered
3. *i* is assigned to *group* 1 if: *π*_1_*f* (**x**_**i**_ | **1**) *> π*_**2**_**f** (**x**_**i**_ | **2**), the classification rule is usually presented in the following form

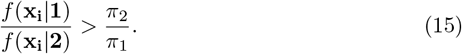 Regarding a priori probabilities, they are usually estimated based on the proportion that each group represents in the sample.
4. If equal a priori probabilities are also assumed, *i* is assigned to *group* 1 if:

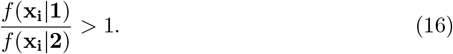

Considering different costs and prior probabilities for each group, *i* is assigned to *group* 1 if: 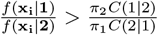 where *C*(1|2) is the cost of classifying a unit in group 1 when it comes from group 2 and *C*(2 | 1) is the cost of classifying a unit as 2 when it comes from group 1.

### Discriminant Function for Linear Discriminant Analysis

The classification rules from which the linear discriminant function is derived are hypothesized as follows:

- Joint normal distribution in each group.
- Equal variance and covariance matrix (Σ_1_ = Σ_2_ = … = Σ_*k*_).

According to what is stated in maximizing *P* (*i* ∈ *g*|*x* = *x*_*i*_) is equivalent to maximizing 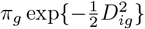. Applying logarithm and replacing *µ* with 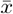 and Σ with *S*:

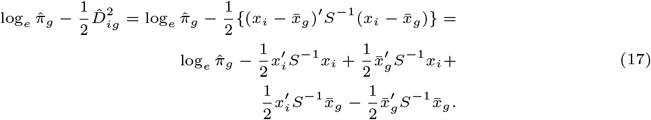

The *“score”* of observation *i* in the **linear discriminant function** estimated for *group g* is:

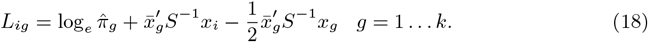

or otherwise the discriminant function is

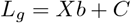

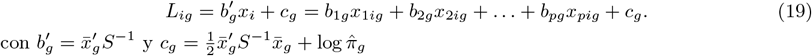

The estimated *classification function* of observation *i* in *group g* is:

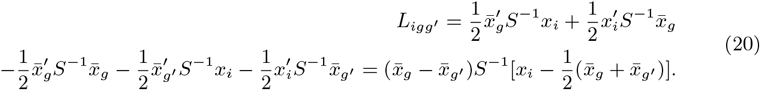

The classification region is

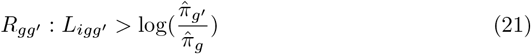

In the case of equal a priori probabilities the rule reduces to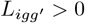. Another way to put it is: observation *i* is classified in the group *g* if 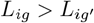 *forall g*^*′*^≠ *g*.

### Logistic Regression Model

The response variable *Y*_*i*_ is a *Bernoulli random variable*, with possible outcomes: *success, failure* encoded as {0, 1}, probability distribution:

*P* (*Y*_*i*_ = 1) = *π*_*i*_, *P* (*Y*_*i*_ = 0) = 1 *− π*_*i*_ and expected value, [19]. ^1^: *E*(*Y*_*i*_|*X*) = *π*_*i*_.

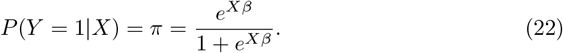

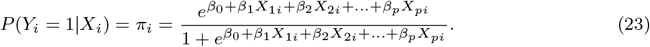

The model proposed in (23) can be linearized by performing the following transformation:

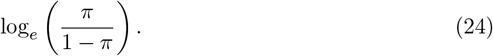

This transformation is called a **Logit transformation**. The quotient 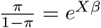 is what is called **odds**, which is a probability ratio, the probability of success over the probability of failure, and it is considered a “measure of risk”. For example, when it is said that the odds are 4 to 1 (odds=4) that a person *x* will suffer from a disease, it is equivalent to saying that the probability of disease is 0.8.

Predicting the value of the response variable based on certain values of other explanatory variables involves determining the critical value above which the estimates of the expected value imply a value of 1 for the response variable. Large values of *π*_*i*_ imply *Y*_*i*_ = 1 while small values of *π*_*i*_ imply *Y*_*i*_ = 0, so it is a problem for the researcher to determine when an estimate is considered a very large or very small value.

### K-neighbor methods

Another binary classifier that can be used and that, unlike AD and ARL, is what is called the *k* nearest neighbors (kNN), which is non-parametric, very easy to implement and based on a very intuitive notion to understand and is based on the Bayesian classifier, which as stated by [21], is the one with the lowest error rate, on average. The Bayesian classifier assigns each observation to the most probable class, conditioned on its predictor values, that is, a test observation, conditioned by the predictor vector *x*_0_ to the class *j* where it is verified

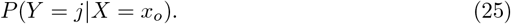

is maximum. For the binary classifier case, the Bayesian classifier that if there are 2 classes *g*_1_ and *g*_2_, *P* (*Y* = 1 | *X* = *x*_0_) *≤* 0.5 ends up classifying in *g*_1_.

Fig shows the *Bayesian decision boundary* for simulated data, where values that fall on the orange side of the boundary will be assigned the orange class

The Bayes classifier will always choose the class for which (25) is largest, so the error rate will be 1 *− max*_*j*_*Pr*(*Y* = *j* | *X* = *x*_0_) at *X* = *x*_0_, [21] state that the *global Bayesian error rate* would be given by:

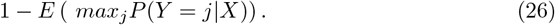

which should be interpreted as the average expected probability value over all possible values of *X*.

As an idea, this seems like something that could work very well; however, it is very difficult to know the conditional distribution of *Y* | *X* for real data. For this reason, one alternative is to estimate the conditional distribution and classify a given observation into the group with the *highest estimated probability*. One method that works with this principle is the *k nearest neighbors*, kNN.

Taking an integer *k* and an observation *x*_0_, the kNN identifies the *k* points closest to *x*_0_ and estimates the conditional probability for cluster *j* as the fraction of the nearest neighbor set *N*_0_, where the answer is *j*

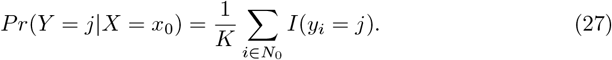

Therefore, the classifier’s performance will depend on the buffer size *k* at each point, where for the same data in Figure 1, changing *k* changes the decision boundary in solid black for the same Bayesian decision boundary in violet dotted line. It can be seen that the change in *k* is important since for *k* = 1 the boundary adapts locally with a *low bias* but *large variance*, while when *k* = 100 the classifier has the inverted properties, and the boundary becomes linear.

**Fig 1.**
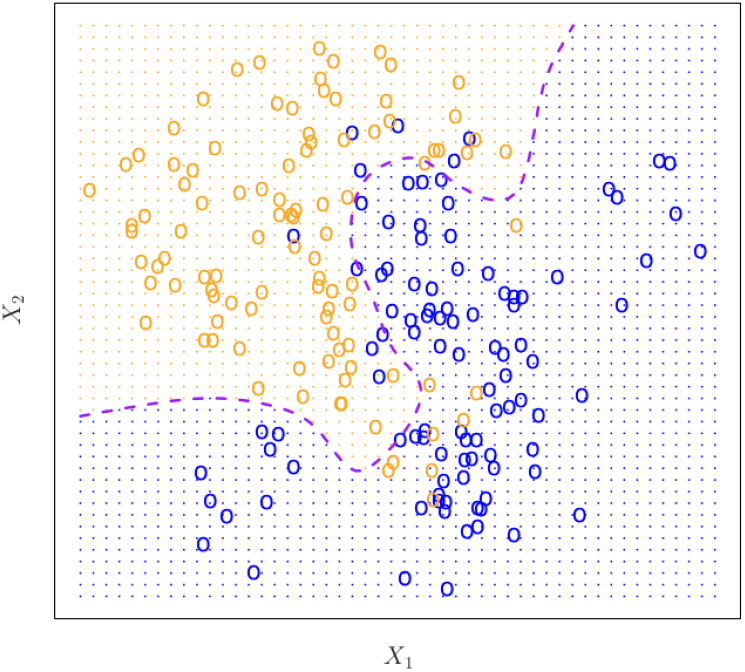
Example of a Bayesian Classifier with 2 groups and 2 variables (taken from An introduction to statistical learning: with applications in R, page 38).

**Fig 2.**
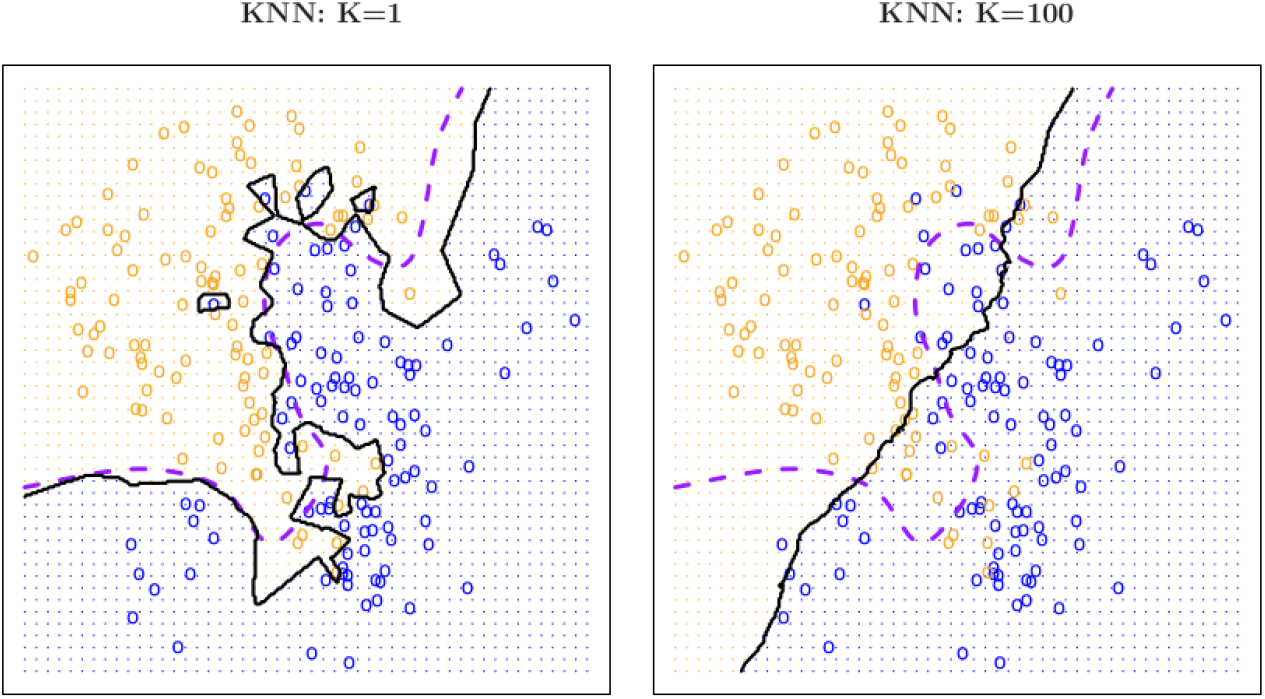
Example of a Bayesian Classifier with 2 groups and 2 variables (extracted from *An introduction to statistical learning* : *with applications in R*, page 41).

**Fig 3.**
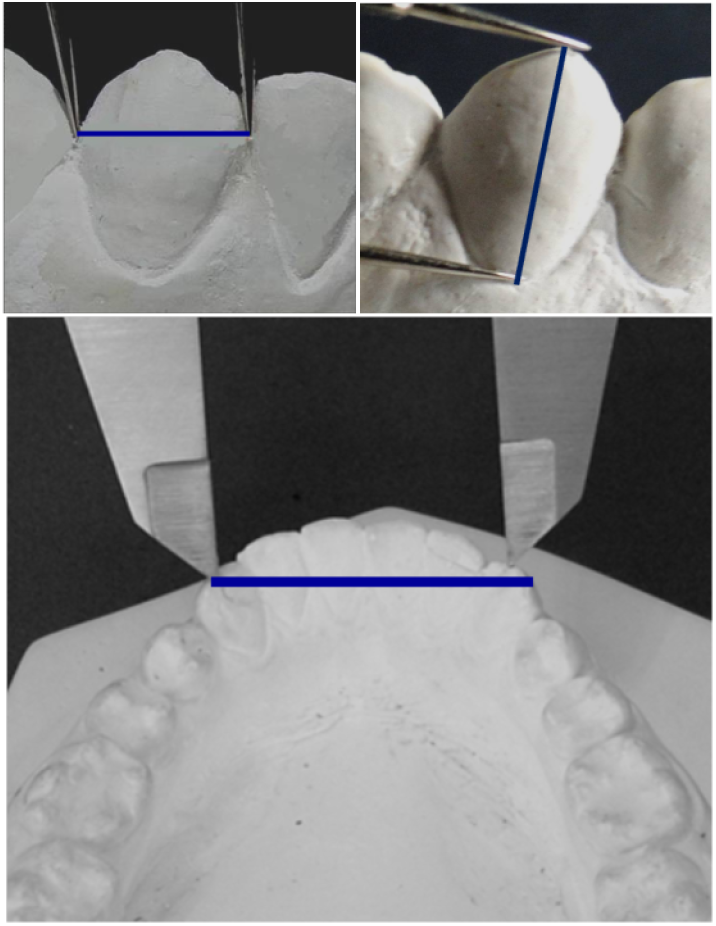
Measures considered in the study (own elaboration).

**Fig 4.**
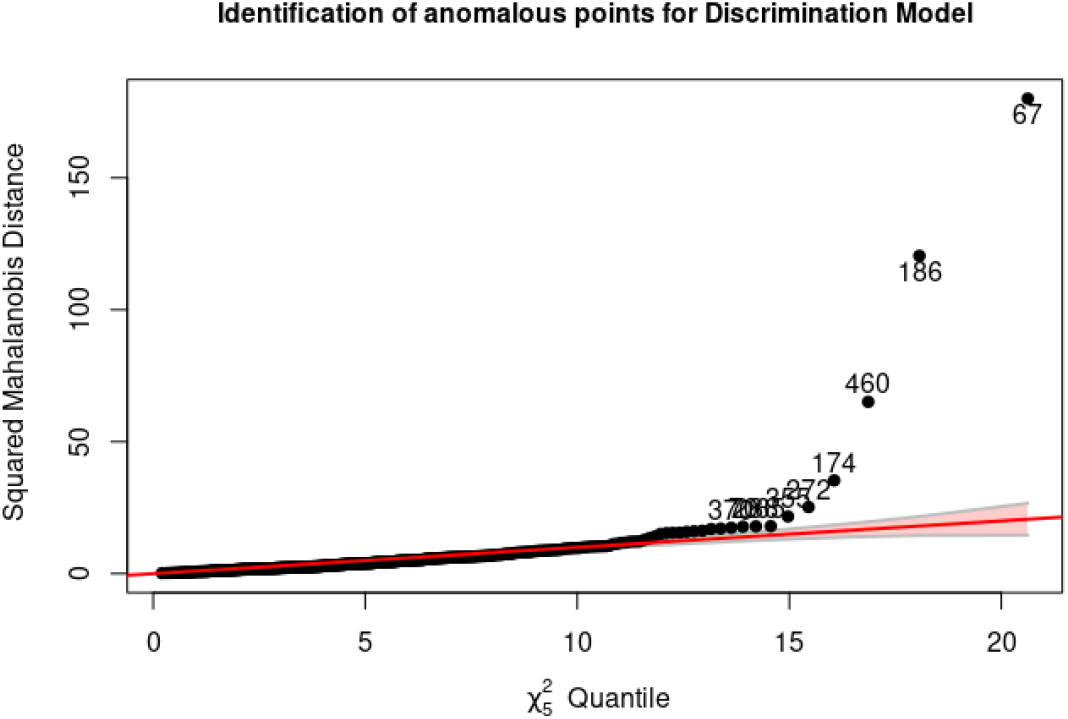
Multivariate anomalous observations using Mahalanobis distance, with complete data.

### Confusion matrices and performance metrics

Usually, as a way of measuring the performance of the different classifiers, in this case binary, what is called *confusion matrix* is used, which allows to see different dimensions of the success capacity for each one, [22], [23], [24].

For this, as a result of the prediction, a table with the following structure is created

From Table 1, a series of metrics are developed that allow the classifier’s different performance characteristics to be evaluated on this data.

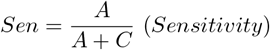

**Table 1.** Confusion matrix.

|  | Reference |  |
| --- | --- | --- |
| Prediction | Event | Not event |
| Event | A | B |
| Not evento | C | D |

**Table 2.** Odontometric measurements by sex.

| Medidas | Statistic | M (N=264) | F (N=260) | p |
| --- | --- | --- | --- | --- |
| DMDd | $\bar{x} \pm DE$ | $7.1 \pm 0.5$ | $6.7 \pm 0.5$ | $< .001$ |
| AGId | $\bar{x} \pm DE$ | $9.3 \pm 1.1$ | $8.9 \pm 0.9$ | $< .001$ |
| DMDi | $\bar{x} \pm DE$ | $7.1 \pm 0.5$ | $6.7 \pm 0.5$ | $< .001$ |
| AGIi | $\bar{x} \pm DE$ | $9.3 \pm 1.1$ | $9.0 \pm 0.9$ | $< .001$ |
| DIC | $\bar{x} \pm DE$ | $26.5 \pm 2.4$ | $25.5 \pm 2.0$ | $< .001$ |

In this case, *sensitivity* measures the model’s ability to detect true events, which in the case of a health condition (patient), i.e., the ability to detect the vast majority of patients.

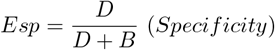

In the case of *specificity*, it is the true negative rate, i.e., the model’s ability to detect healthy individuals in the face of a health condition.

Therefore, it is desirable to have both metrics with maximum values, which is not possible since they have a negative correlation. A change is any of the metrics; it involves changing the cutoff threshold, which changes the prediction, and in doing so, the confussion matrix is reconfigured.

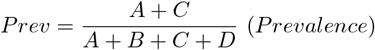

Continuing with the possible situation of the event being a case of disease, it is also important to know the fraction of people observed to have the condition of the event, which is known as prevalence, a term derived from epidemiology.

The positive and negative predictive values (Pred Value (+) and Pred Value (-)) respectively) are the proportions of positive and negative results generated by a classifier that are actually true positives and true negatives, respectively. High values for both metrics can be interpreted as an indicator of the accuracy of the result generated by the classifier. The Predictive Value (+) and Predictive Value (-) are not intrinsic to the classifier, as is the case with the true positive rate and the true negative rate, since the Predictive Values depend on the prevalence, as shown in the formula. It should be noted that the Predictive Value (+) and Predictive Value (-), like the NPV, can be derived using Bayes’ theorem. A first expression for Pred Value (+) is

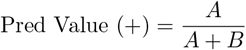

This can be rewritten through sensitivity, specificity, and prevalence

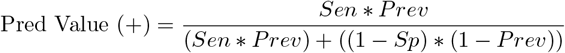

For the case of Pred Value (-)

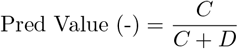

Using sensitivity, specificity, and prevalence for Pred Value (-) we have

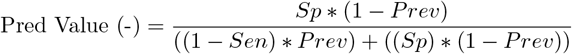

The Detection Rate can be viewed as the classifier’s ability to detect as events, cases that actually constitute events, out of the total number of cases:

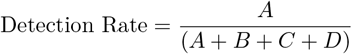

While the Detection Prevalence can be viewed as the proportion of events that the classifier classifies as such out of the total number of cases:

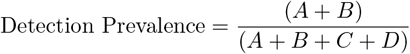

Balanced Precision is an important metric as it shows the classifier’s performance by averaging the ability to detect the true positive rate and the true negative rate.

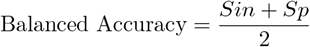

There are two other metrics that are also widely used in clinical diagnostic tests: the positive likelihood ratio LR(+) (in English, LR(+)).

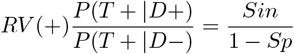

where the expression *P* (*T* + | *D*+) can be seen as the probability that a given person with the disease will be classified by the classifier as sick (since it is an event, it will be classified as an event). This can be expressed through sensitivity and specificity.

The same can be said for the positive likelihood ratio LR(-) (in English, English LR(-))

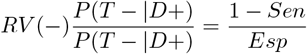

Combining both likelihood ratios produces another metric called ORD (diagnostic odds ratio), which measures the effectiveness of the classifier and expresses the relationship between the probabilities of the classifier being positive, if the subject is really positive (has the disease) in relation to the probabilities of the classifier being positive, if the subject does not have the disease.

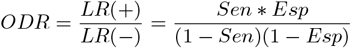

In the statistical analysis of information retrieval and binary classification systems, the F-score or F-measure is a measure of predictive performance. It is calculated from the test’s accuracy and sensitivity, where accuracy is the number of true positive results divided by the number of all samples predicted to be positive, including those not correctly identified.

The *F*_1_ metric is the harmonic mean of the precision (which is the Pred Value (+)) and the sensitivity (called recall). Therefore, it symmetrically represents both precision and recall in a single metric. For other *β* values, differential weights are applied, valuing one or recall more than the other.

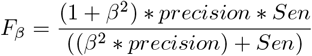

This expression has a direct link to the confusion matrix seen from the *Type I error* and *Type II error* in the context of hypothesis testing.

## Materials

In this work, only the measurements of 524 lower plaster models (286 male subjects and 238 females) from patients treated at an orthodontic clinic in Montevideo, Uruguay, are taken into account. The mesiodistal diameter (width), gingivo-incisal height of the 4 canines, and the intercanine distance were measured for sex identification based on odontometric measurements and their ratios, from a larger study published in [7, 20]. To evaluate the performance of the binary classifiers presented in previous sections, the metrics presented in section about Confusion matrices and performance metrics are used. The work is implemented in R language [25] and used several libraries like MASS [18], klar [26], heplots [27], rsample [28], class [18] libraries. For the graphics, ggplot2 [29], GGally [30] are used, and for the different metrics, caret [24]. The computational code is available at https://osf.io/javru/

## Results and Discussion

After evaluating which measures had anomalous values, the sample is reduced to 511 by removing the values that have *DMDd ≥* 8.5 or *AGId ≥* 11.5 and also for *DMDi ≥* 8.5 or *AGIi ≥* 11.5. Although in Figure 5 there are almost 30 anomalous values, which are outside the confidence band, but in Table 3, they do not essentially change the mean values but which by themselves can be ”lever” points.

**Table 3.** Odontometric measurements by sex without outliers.

| Measures | Statistic | M (N=255) | F (N=256) | p |
| --- | --- | --- | --- | --- |
| DMDd | $\bar{x} \pm DE$ | $7.1 \pm 0.4$ | $6.7 \pm 0.4$ | < .001 |
| AGId | $\bar{x} \pm DE$ | $9.2 \pm 1.0$ | $8.9 \pm 0.9$ | < .001 |
| DMDi | $\bar{x} \pm DE$ | $7.1 \pm 0.5$ | $6.7 \pm 0.4$ | < .001 |
| AGIi | $\bar{x} \pm DE$ | $9.3 \pm 1.0$ | $9.0 \pm 0.9$ | < .001 |
| DIC | $\bar{x} \pm DE$ | $26.4 \pm 2.3$ | $25.5 \pm 2.0$ | < .001 |

**Fig 5.**
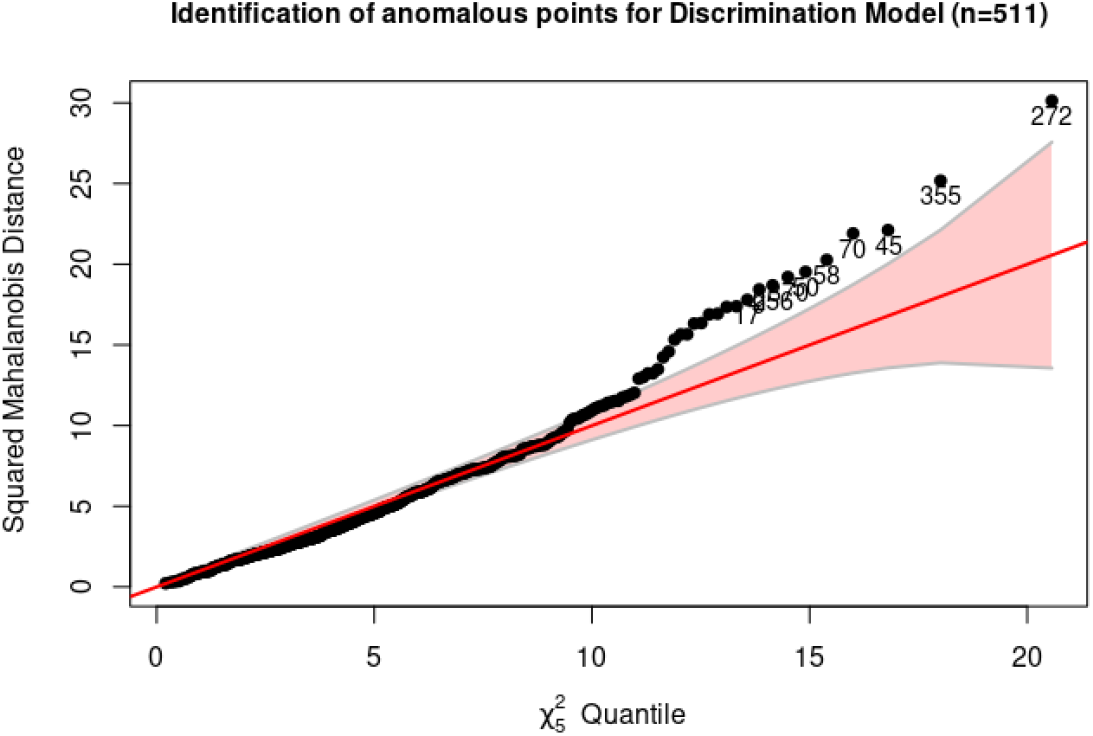
Multivariate anomalous observations using Mahalanobis distance.

With the data filtered, the territorial maps are presented using the 2-by-2 variables and considering linear and quadratic boundaries. These maps show areas with a significant mix of observations that are misclassified and where there is significant overlap. For the AD, ARL, and kNN methods, it is proposed to work with a 75% learning sample and a 25% testing sample.

Looking at the results obtained in Table 4, only one measurement remains as part of the model, and it is one-sided, after applying a sequential ANOVA to remove variables. This means that, from a practical point of view, having a model that only uses canine diameter or only measurements from one side does not make sense for forensic specialists, which is why it is decided to develop two models, only with measurements for each side.

**Table 4.** Logistic Regression without outliers.

| Measures | M (N=255) | F (N=256) | OR (multivariate) |
| --- | --- | --- | --- |
| DMDd | 7.1 ± 0.4 | 6.7 ± 0.4 | 0.25 (0.11-0.54, < .001) |
| AGId | 9.2 ± 1.0 | 8.9 ± 0.9 | 0.78 (0.53-1.17, *p* = .234) |
| DMDi | 7.1 ± 0.5 | 6.7 ± 0.4 | 0.55 (0.27-1.15, *p* = .114) |
| AGIi | 9.3 ± 1.0 | 9.0 ± 0.9 | 1.16 (0.77-1.75, *p* = .468) |
| DIC | 26.4 ± 2.3 | 25.5 ± 2.0 | 0.92 (0.84-1.01, *p* = .088) |

**Table 5.**
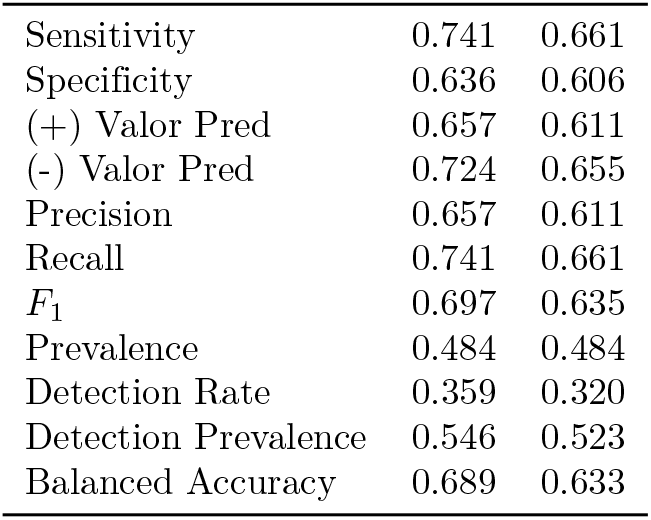
Metrics for ADL and ADC for testing sample.

|  |  |  |
| --- | --- | --- |
| Sensitivity | 0.741 | 0.661 |
| Specificity | 0.636 | 0.606 |
| (+) Valor Pred | 0.657 | 0.611 |
| (-) Valor Pred | 0.724 | 0.655 |
| Precision | 0.657 | 0.611 |
| Recall | 0.741 | 0.661 |
| *F*<sub>1</sub> | 0.697 | 0.635 |
| Prevalence | 0.484 | 0.484 |
| Detection Rate | 0.359 | 0.320 |
| Detection Prevalence | 0.546 | 0.523 |
| Balanced Accuracy | 0.689 | 0.633 |

The same learning sample is used and the results in tables 6 and 7 show that the models on each side are still incomplete since they only consider one measure and therefore, to evaluate the dependence of the results on the training sample, it is proposed to see the robustness by resampling *boostrap* with 500 samples, where in each one the data table is split into 10 learning/validation samples by cross-validation (V-Fold Cross-Validation), [24].

**Table 6.** Logistic Regression without Outliers for Right Side.

| Measures | M (N=255) | F (N=256) | OR (multivariate) |
| --- | --- | --- | --- |
| DMDd | 7.1 ± 0.4 | 6.7 ± 0.4 | 0.16 (0.09-0.26, $p < .001$) |
| AGId | 9.2 ± 1.0 | 8.9 ± 0.9 | 0.89 (0.72-1.11, $p = .299$) |
| DIC | 26.4 ± 2.3 | 25.5 ± 2.0 | 0.92 (0.83-1.01, $p = .065$) |

**Table 7.** Logistic Regression without Outliers for Left Side.

| Measures | M (N=255) | F (N=256) | OR (multivariate) |
| --- | --- | --- | --- |
| DMDi | 7.1 ± 0.5 | 6.7 ± 0.4 | 0.20 (0.12-0.32, $p < .001$) |
| AGIi | 9.3 ± 1.0 | 9.0 ± 0.9 | 0.88 (0.71-1.09, $p = .234$) |
| DIC | 26.4 ± 2.3 | 25.5 ± 2.0 | 0.90 (0.82-0.99, $p = .022$) |

**Table 8.** Metrics for *k* = 3 a *k* = 11.

| $K$ | 3 | 5 | 7 | 9 | 11 |
| --- | --- | --- | --- | --- | --- |
| Sensitivity | 0.532 | 0.532 | 0.645 | 0.677 | 0.661 |
| Specificity | 0.606 | 0.591 | 0.561 | 0.606 | **0.652** |
| (+) Valor Pred | 0.559 | 0.550 | 0.580 | 0.618 | 0.641 |
| (-) Valor Pred | 0.580 | 0.574 | 0.627 | 0.667 | 0.672 |
| Precision | 0.559 | 0.550 | 0.580 | 0.618 | 0.641 |
| Recall | 0.532 | 0.532 | 0.645 | 0.677 | 0.661 |
| F1 | 0.545 | 0.541 | 0.611 | 0.646 | 0.651 |
| Prevalence | 0.484 | 0.484 | 0.484 | 0.484 | 0.484 |
| Detection Rate | 0.258 | 0.258 | 0.312 | 0.328 | 0.320 |
| Detection Prevalence | 0.461 | 0.469 | 0.539 | 0.531 | 0.500 |
| Balanced Accuracy | 0.569 | 0.562 | 0.603 | 0.642 | 0.656 |

**Table 9.** Metrics for *k* = 13 a *k* = 21.

| $K$ | 13 | 15 | 17 | 19 | 21 |
| --- | --- | --- | --- | --- | --- |
| Sensitivity | 0.661 | 0.694 | 0.661 | 0.710 | **0.726** |
| Specificity | 0.636 | 0.636 | 0.621 | 0.636 | 0.636 |
| (+) Valor Pred | 0.631 | 0.642 | 0.621 | 0.647 | **0.652** |
| (-) Valor Pred | 0.667 | 0.689 | 0.661 | 0.700 | **0.712** |
| Precision | 0.631 | 0.642 | 0.621 | 0.647 | **0.652** |
| Recall | 0.661 | 0.694 | 0.661 | 0.710 | **0.726** |
| F1 | 0.646 | 0.667 | 0.641 | 0.677 | **0.687** |
| Prevalencia | 0.484 | 0.484 | 0.484 | 0.484 | 0.484 |
| Detection Rate | 0.320 | 0.336 | 0.320 | 0.344 | **0.352** |
| Detection Prevalence | 0.508 | 0.523 | 0.516 | 0.531 | **0.539** |
| Balanced Accuracy | 0.649 | 0.665 | 0.641 | 0.673 | **0.681** |

Both figures 6 and 7 show that the ARL only considers a single measure, which is why it is ruled out as a parametric model, as it only considers a single measure in each model, even though they may have good performance in classification metrics.

**Fig 6.**
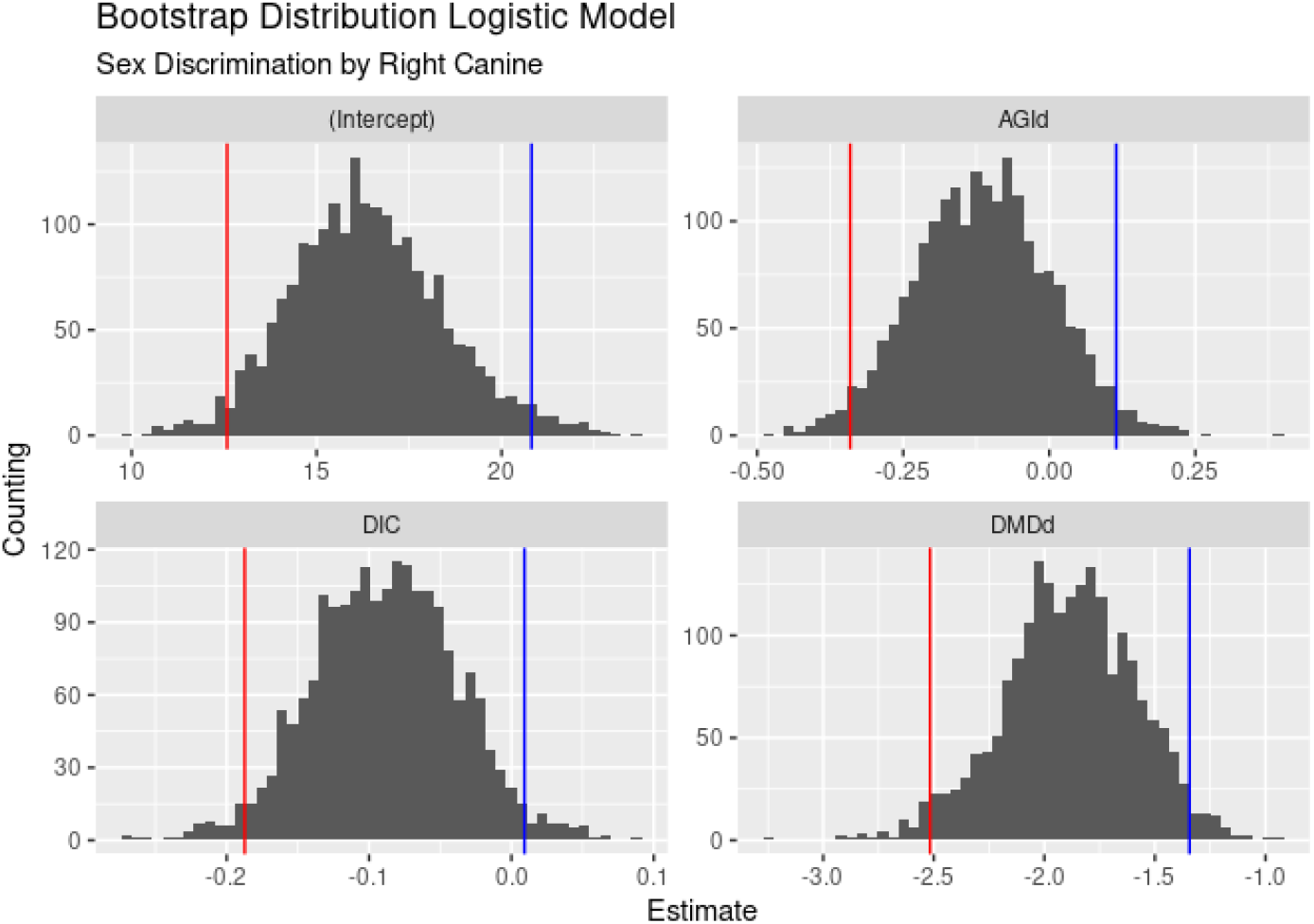
Bootstrap Distribution Logistic Model, Right side, 500 samples.

**Fig 7.**
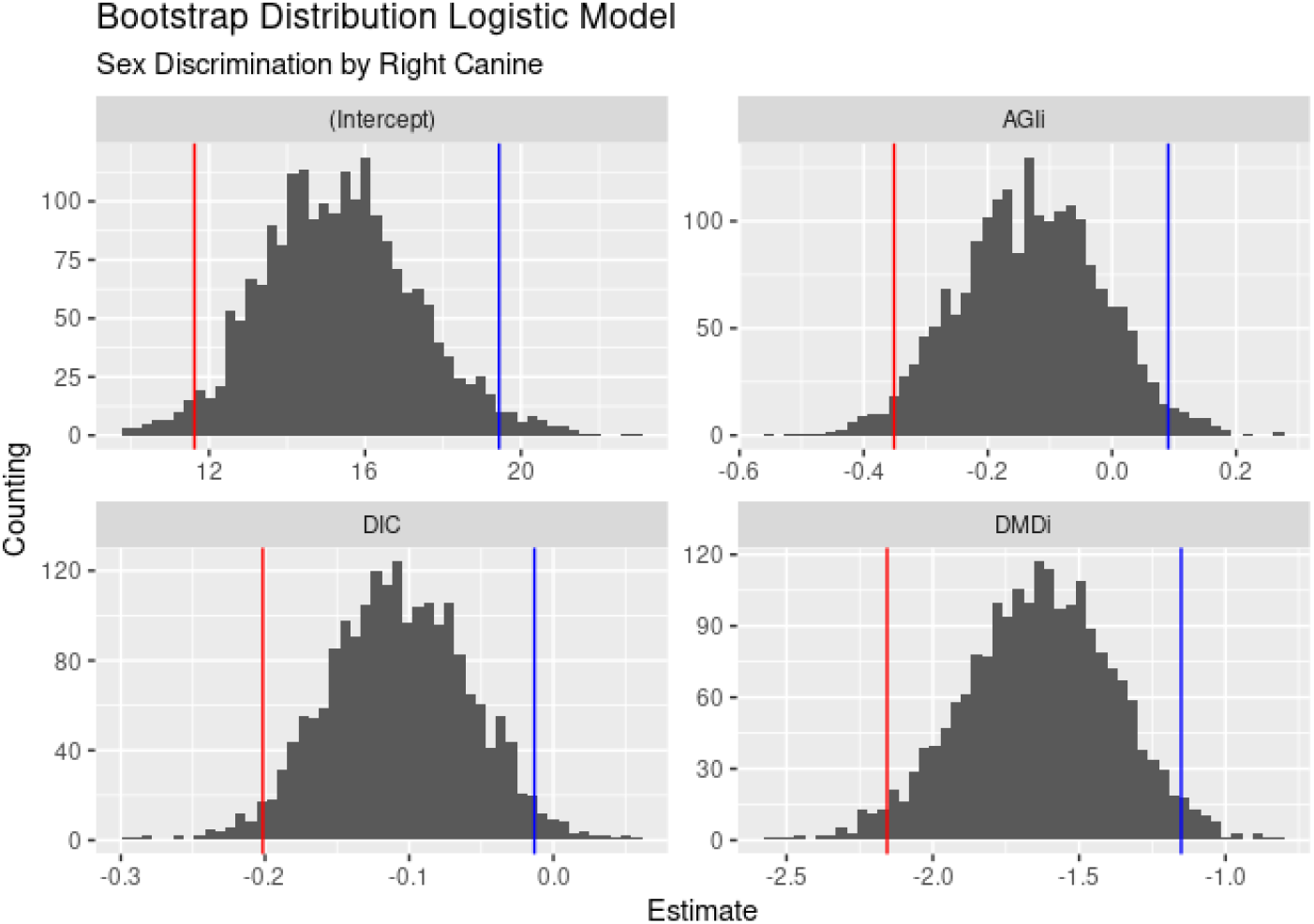
Bootstrap Distribution Logistic Model, Left side, 500 samples.

**Fig 8.**
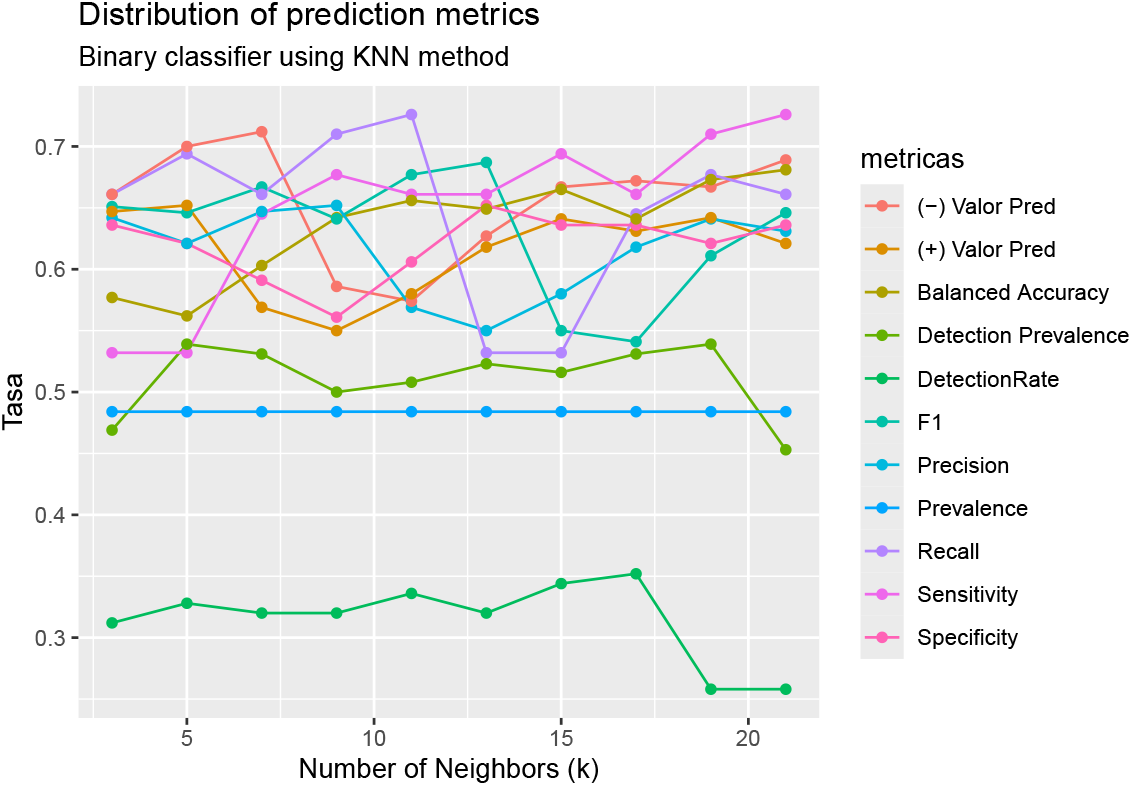
Performance of the classification model.

Finally, for comparison purposes, the *k − neighbors* method is applied to the same training sample used for the ADL and ADC, changing the parameter *k* to see how the metrics vary.

When comparing the three classifiers(*AD, ARL*, and *kNN*), it is observed that the data present anomalous values that must be filtered out.However, despite this, multinormality is not verified for AD, which should justify its discarding, despite its good performance as a classifier. When using an alternative, also parametric, classifier such as ARL, without the restrictions of distribution in the two groups, the measurements appear to be correlated, which prevents obtaining a model with all five variables. By discarding nonsignificant variables, two models are chosen that consider the two dimensions of each canine (height and diameter), considering each side.However, the idea persists that the only relevant measurement is the mediodistal diameter for both sides, which, strictly speaking, makes it unusable for forensic specialists. Bootstrap resampling corroborates that this is a characteristic of the data and that a potential learning sample does not depend on it. Finally, the third binary classifier used is the kNN, using several values of *k*, where it can be observed that since the reference for classification is the male sex, the sensitivity (the rate of correctly classified men) vs. the specificity (the rate of correctly classified women) changes, showing a slight bias depending on *k*, with a behavior that is not monotonous and that if compared with the AD, the bias is maintained and, in addition, for the ADC the metrics are impoverished.

## Conclusion

The conclusions so far show that the classifiers’ performance does not differ drastically when changing, and in particular for the ARL, the fact that only one measure is significant suggests that another attribute should be considered, which was not used because it is a qualitative attribute: crowding (crowded teeth) or diastema (spaced teeth). This attribute was not used to allow for binary classifiers for any type of teeth, and not if those data are removed, which would lead to binary classifiers for normal teeth.

For this reason, three paths to follow are proposed as a continuation of the work.

- Filter the data for teeth without crowding or diastema pathologies and evaluate the three proposed classifiers.
- For the case of the ARL, not only evaluate the relevance of joint or side-by-side models using the two measurements of each canine, but also the performance of the classification metrics (which was not done in this work).
- For crowded or diastema teeth, evaluate the three classifiers separately.
- For all teeth, test an ARL considering the crowding variable as a regressor, which would perhaps allow for a global model, but which would not be comparable with the other classifiers.

Furthermore, as alternatives to testing other classifiers, it is proposed to see how support vector machines that only work with quantitative regressor variables work, [21, cap 9], and classification trees that support any type of variable, being a non-parametric method that makes it comparable to ARL and that also allows extension through techniques such as random forests and *boosting* and *bagging* methods, [21, cap 8].

## Data Availability

https://doi.org/10.60895/redata/JIU5UG

## Supporting information

**S1 Fig. Example of a Bayesian Classifier with 2 groups and 2 variables (taken from An introduction to statistical learning: with applications in R, page 38)**.

**S2 Fig. Example of a Bayesian Classifier with 2 groups and 2 variables (extracted from *An introduction to statistical learning* : *with applications in R*, page 41)**.

**S3 Fig. Measures considered in the study (own elaboration)**.

**S4 Fig. Multivariate anomalous observations using Mahalanobis distance, with complete data**.

**S5 Fig. Multivariate anomalous observations using Mahalanobis distance, with complete data**.

**S6 Fig. Multivariate anomalous observations using Mahalanobis distance**.

**S7 Fig. Bootstrap Distribution Logistic Model, Left side, 500 samples**.

**S8 Fig. Performance of the classification model**.

**S1 Table. Confusion matrix**.

**S2 Table. Odontometric measurements by sex**.

**S3 Table. Odontometric measurements by sex without outliers**.

**S4 Table. Logistic Regression without outliers**.

**S5 Table. Metrics for ADL and ADC for testing sample**.

**S6 Table. Logistic Regression without Outliers for Right Side**.

**S7 Table. Logistic Regression without Outliers for Left Side**.

**S8 Table. Metrics for k = 3 a k = 11**.

**S9 Table. Metrics for k = 13 a k = 21**.

## Acknowledgments

Not applicable

## Footnotes

1 For the purpose of simplifying the notation in the text, *π*_*i*_ = *π*(*X*_*i*_) = *E*(*Y*_*i*_|*X*_*i*_) will be used

## Notes

### Competing Interest Statement

The authors have declared no competing interest.

### Funding Statement

This study did not receive any funding

### Author Declarations

It is hereby noted that the Ethics Committee of the Facultad de Odontología - Universidad de la República Uruguay, in its meeting held on August 1, 2013, resolved to approve the research project entitled "Dental Anthropology: Dentist Participation," submitted by Dr. Veronica Gargano and Dr. Alicia Picapedra, file no. 091900-000098-13. The Ethics Committee members are Dr. Ernesto Borgia (Chair), Dra. Sivana Blanco, Dr. Jorge Gutierrez, Dr. Carolina Seade, Dr. Felipe Luzardo, and Social Worker Ema Menoni.

